# High Mortality among Older Patients Hospitalized with COVID-19 during the First Pandemic Wave

**DOI:** 10.1101/2022.06.16.22276514

**Authors:** Russell R Kempker, Paulina A Rebolledo, Francois Rollin, Saumya Gurbani, Marcos C. Schechter, David Wilhoite, Sherri N. Bogard, Stacey Watkins, Aarti Duggal, Nova John, Malavika Kapuria, Charles Terry, Philip Yang, Gordon Dale, Ariana Mora, Jessica Preslar, Kaitlin Sandor, Yun F (Wayne) Wang, Michael H Woodworth, Jordan A Kempker

## Abstract

**Background:** Understanding the local epidemiology, including mortality, of COVID-19 is important for guiding optimal mitigation strategies such as vaccine implementation, need for study of more effective treatment, and redoubling of focused infection control measures.

**Methods:** A retrospective observational cohort study design was utilized. We included adult patients diagnosed in the hospital or emergency department with COVID-19 from March 8, 2020 through May 17, 2020 at Grady Memorial Hospital (Atlanta, GA). Medical chart data abstraction was performed to collect clinical, laboratory and outcome data. Death, defined as inpatient mortality or discharge to hospice, was the primary outcome.

**Results:** Among 360 persons with laboratory-confirmed COVID-19, 50% were ≥ 60 years, and most (80%) were Black and had a BMI ≥25 kg/m^2^ (64%). A total of 53 patients (15%) had an outcome of death with the majority (n=46, 88%) occurring in persons ≥ 60 years. Persons ≥ 60 years were less likely to have typical COVID-19 symptoms while more likely to have multiple comorbidities, multifocal pneumonia, and to be admitted to intensive care. The death rate was 27% among persons ≥60 years versus 4% in those <60 years (p<.01). Furthermore, most deaths (n=40, 75%) occurred among residents of long-term care facilities (LCFs).

**Conclusions:** We describe early COVID-19 cases among predominantly Black and older patients from a single center safety net hospital. COVID-19 related mortality occurred predominantly among older patients from LCFs highlighting the need for improved preparedness and supporting prioritization of vaccination efforts in such settings.

## INTRODUCTION

The novel coronavirus (COVID-19) pandemic took the world by surprise in early 2020 and has led to a staggering death toll during the subsequent year with no immediate end in sight. Researchers quickly mobilized to combat the pandemic, turning COVID-19 into the predominant and persistent focus of the scientific and medical communities.^1^ Unprecedented breakthroughs in therapeutics and vaccines have provided hope.^2, 3^ However, health inequities, vaccine uptake and distribution challenges, human behavior, emerging viral variants, and other challenges will likely contribute to the continued circulation of SARS-CoV-2.^4, 5^ Massive epidemiological efforts focused on COVID-19 have revealed that populations affected and clinical outcomes differ by time and place stressing the importance of understanding the pandemic at a local level. ^6, 7^

Utilizing available estimates, the United States has had one of the highest prevalences of COVID-19 cases and COVID-19 related deaths in the world. As of March 18, 2021 the U.S. accounted for ∼24% (∼29.3 million) of all COVID-19 cases worldwide as well as ∼20% (∼531,855) of all deaths.^7, 8^ While COVID-19 has affected all strata of the population, increasing age and residence in a long-term care facility (LCF) were identified early on as risk factors for COVID-19 acquisition and severe disease. One of the first reports from the U.S. detailed the fast spread of COVID-19 among LCFs in Seattle highlighting the importance of preventive measures in such settings.^9^ A subsequent report from 18 states found that >50% of all their COVID-19 related deaths occurred among persons living in LCFs and estimated that > 100,000 COVID-19 deaths in the U.S. have occurred among LCFs residents.^10^ Furthermore, while recent data has implicated COVID-19 as the leading cause of death in the U.S., the highest death rates were seen in persons > 65 years.^11^

The purpose of our study was to evaluate the clinical characteristics of patients with COVID-19 at an urban, safety-net hospital in the South during the first wave of the pandemic and to identify patient factors associated with mortality from COVID-19. As the COVID-19 pandemic has evolved to include broader segments of the population, additional data to identify those most at risk of severe disease in certain settings is important to guide optimal use of limited resources now and in the future. We expect our data will inform ongoing surveillance and prevention efforts in the South and potentially provide valuable information to encourage high risk groups including those with higher rates of vaccine hesitancy to be immunized.^12^

## METHODS

### Study Design and Participants

A retrospective observational cohort study design was utilized. We included adult patients diagnosed with COVID-19 in either the hospital or emergency room from March 8, 2020 through May 17, 2020 at Grady Memorial Hospital. Grady Memorial Hospital is an 800-bed safety-net hospital in Atlanta, Georgia which diagnosed its first case of COVID-19 on March 8, 2020. Of note during the study period, the Grady nursing to patient ration remained between 1:1 to 1:3 in the intensive care unit. A positive polymerase chain reaction (PCR) assay for SARS-CoV-2 was required for inclusion. During our study period, nasopharyngeal swab samples were collected for all persons under investigation and analyzed with the Abbott Laboratories *m*2000 RealTime (Lake Bluff, IL) system. Patients who were diagnosed in the outpatient setting without an admission order were excluded. Additionally, participants with their first SARS-CoV-2 PCR test performed ≥5 days after hospitalization were excluded. The study was approved by the Emory University Institutional Review Board and the Grady Memorial Hospital Research Oversight Committee.

### Data Management

COVID-19 surveillance data was obtained from Grady Memorial Hospital EPIC electronic medical record (EMR) system (Epic Systems Corporation, Verona, WI) as well as from the microbiology laboratory. Utilizing the EPIC Clinical Workbench Reporting feature, we exported a list of patients who had received a SARS-CoV-2 test. Additionally, a list of patients admitted to the hospital who had a SARS-CoV-2 test was generated from the microbiology laboratory. Demographic information including age, sex, and race were included in the data pull when available. Data were exported from both methods on a weekly basis. An R script was created in RStudio (version 1.2.5, RStudio Inc., 2020) that cross-referenced patients from the two data sets using the medical record number and updated a cumulative R dataset weekly. The primary variable for assessing disease prevalence was the SARS-CoV-2 test result, and the date the sample was received by the laboratory was used as the time point. If a patient had multiple positive SARS-CoV2 test results over the collection period, only the first result and time were included in analyses. At the end of the collection period, the R data were uploaded into an HIPAA compliant online Research Electronic Data Capture (REDCap) database^13^ hosted by Emory University. Utilizing the list of patients with a positive SARS-CoV-2 test generated above, an electronic medical chart data abstraction was performed for each participant and data (patient demographics, medical history, symptoms, laboratory values, radiology results, treatment and clinical outcomes) was entered into the REDCap database.

### Data Analysis

Data analyses were performed using RStudio (version 3.6.3, R Core Team, 2020). Death was the main outcome of interest and was defined as inpatient mortality or discharge to hospice. Older patients were defined by age ≥60 years. Sequential organ failure assessment (SOFA) and quick SOFA (qSOFA) scores were calculated for each patient with simplified vasopressor categories using a custom R package (https://github.com/michaelwoodworth/sepsisscores).^14^ Bivariate analysis was performed to compare characteristics (demographics, comorbidities, treatment received and laboratory and radiology results) between patients by vital status and by age category.

## RESULTS

A total of 4049 participants had a SARS-CoV2 PCR test performed during the study period including 34.6% who were ≥60 years and 70.9% who were Black or African American. A total of 360 (8.9%) had a positive test. Among the 360 patients with COVID-19, 50% were ≥60 years, and 80% were Black and 64% had a body mass index ≥25 kg/m^2^. Regarding place of residence prior to admission, 106 patients (29%) resided in a LCF (Table 1). The most common comorbidity was hypertension (66%) followed by diabetes mellitus (31%) (Table 1). The majority of patients (67%) had an abnormal chest radiograph with bilateral opacities being the most common finding (45%). Almost one quarter (24%) of patients were admitted to either intermediate or intensive care and 16% of patients received mechanical ventilation during their hospitalization. Only 14 patients (4%) received remdesivir as it only became available near the end of the study period through participation in a clinical trial.

**Table 1.** Demographic and Medical Characteristics of Hospitalized Patients with COVID-19 by Mortality Status

|  | Alive (N=307) | Died (N=53) | Total (N=360) | p |
| --- | --- | --- | --- | --- |
| **Age category, years** |  |  |  | < 0.001 |
| <20 | 7 (2%) | 0 (0%) | 7 (2%) |  |
| 20-39 | 55 (18%) | 0 (0%) | 55 (15%) |  |
| 40-59 | 110 (36%) | 6 (11%) | 116 (32%) |  |
| 60-79 | 116 (38%) | 32 (60%) | 148 (41%) |  |
| >/=80 | 19 (6%) | 15 (28%) | 34 (9%) |  |
| **Female** | 143 (47%) | 16 (30%) | 159 (44%) | 0.026 |
| **Race** |  |  |  | 0.519 |
| Black | 240 (78%) | 46 (87%) | 286 (80%) |  |
| White | 54 (18%) | 6 (11%) | 60 (17%) |  |
| Other | 4 (1%) | 0 (0%) | 4 (1%) |  |
| **Hispanic** | 38 (13%) | 4 (8%) | 42 (12%) | 0.314 |
| **Housing type** |  |  |  | < 0.001 |
| Correctional facility | 14 (5%) | 0 (0%) | 14 (4%) |  |
| Homeless/shelter | 17 (6%) | 0 (0%) | 17 (5%) |  |
| Long term health facility | 7 (2%) | 1 (2%) | 8 (2%) |  |
| Nursing home | 59 (19%) | 39 (74%) | 98 (27%) |  |
| Stable Home | 207 (68%) | 13 (25%) | 220 (61%) |  |
| **BMI (kg/m2) categories** |  |  |  | 0.288 |
| <18.5 | 22 (7%) | 4 (8%) | 26 (7%) |  |
| 18.5-24.9 | 77 (26%) | 20 (38%) | 97 (28%) |  |
| 25-29.9 | 82 (28%) | 13 (25%) | 95 (27%) |  |
| ≥30 | 115 (39%) | 15 (29%) | 130 (37%) |  |
| **Hypertension** | 191 (62%) | 45 (85%) | 236 (66%) | 0.001 |
| **Diabetes mellitus** |  |  |  | 0.004 |
| IDDM | 43 (14%) | 4 (8%) | 47 (13%) |  |
| NIDDM | 53 (17%) | 11 (21%) | 64 (18%) |  |
| **Congestive heart failure** | 42 (14%) | 13 (25%) | 55 (15%) | 0.037 |
| **Coronary artery disease** | 27 (9%) | 5 (10%) | 32 (9%) | 0.853 |
| **Stroke** | 61 (20%) | 26 (49%) | 87 (24%) | < 0.001 |
| **Dementia** | 47 (15%) | 29 (55%) | 76 (21%) | < 0.001 |
| **Chronic kidney disease** |  |  |  | 0.003 |
| Yes | 29 (9%) | 11 (21%) | 40 (11%) |  |
| Yes, on dialysis | 25 (8%) | 1 (2%) | 26 (7%) |  |
| **Cirrhosis** | 3 (1%) | 1 (2%) | 4 (1%) | 0.562 |
| **COPD** | 17 (6%) | 5 (9%) | 22 (6%) | 0.274 |
| **Asthma** | 26 (8%) | 2 (4%) | 28 (8%) | 0.239 |
| **Structural lung disease** | 4 (1%) | 1 (2%) | 5 (1%) | 0.737 |
| **OSA/OHS** | 15 (5%) | 4 (8%) | 19 (5%) | 0.675 |
| **Malignancy** |  |  |  | 0.783 |
| Current | 10 (3%) | 1 (2%) | 11 (3%) |  |
| Prior | 9 (3%) | 1 (2%) | 10 (3%) |  |
| **HIV** | 13 (4%) | 0 (0%) | 13 (4%) | 0.284 |

\*BMI, body mass index; IDDM, insulin dependent diabetes mellitus; NIDDM, non-insulin dependent
diabetes mellitus; COPD, chronic obstructive pulmonary disease; OSA/OHS, obstructive sleep
apnea/obesity hypoventilation syndrome; HIV, human immunodeficiency virus

There were a total of 53 deaths (15%), including 31 with in-hospital death and 22 patients discharged to hospice. The majority of deaths (87%) occurred among patients ≥60 years old and among persons who resided in a LCF prior to admission (76%). No deaths occurred in patients <40 years old. Approximately half of patients admitted to the intensive care (51%) and patients receiving mechanical ventilation (54%) died.

Given the high rate of death in older persons with COVID-19, we next compared characteristics by age category (≥60 vs. < 60 years). Older patients were more likely to have multiple chronic comorbidities including hypertension, congestive heart failure, prior cerebrovascular accident, dementia, and chronic kidney disease (Table 2). Older patients were less likely to have upper respiratory viral symptoms compared to patients < 60 years (including fever, cough, chills and myalgias). The only symptom more common in older patients was altered mental status (46 vs. 8%, p<.01). Multifocal lung lesions were more common on chest x-ray in older patients (21 vs. 11%, p=0.01). Older patients were also more likely to be admitted to either intermediate or intensive care (35 vs. 14%, p<.01) and more likely to receive mechanical ventilation (22 vs. 10%, p<.01). Among patients with COVID-19, a total of 27% older persons compared to 4% (p<.01) of patients < 60 years died during the study period.

**Table 2.**
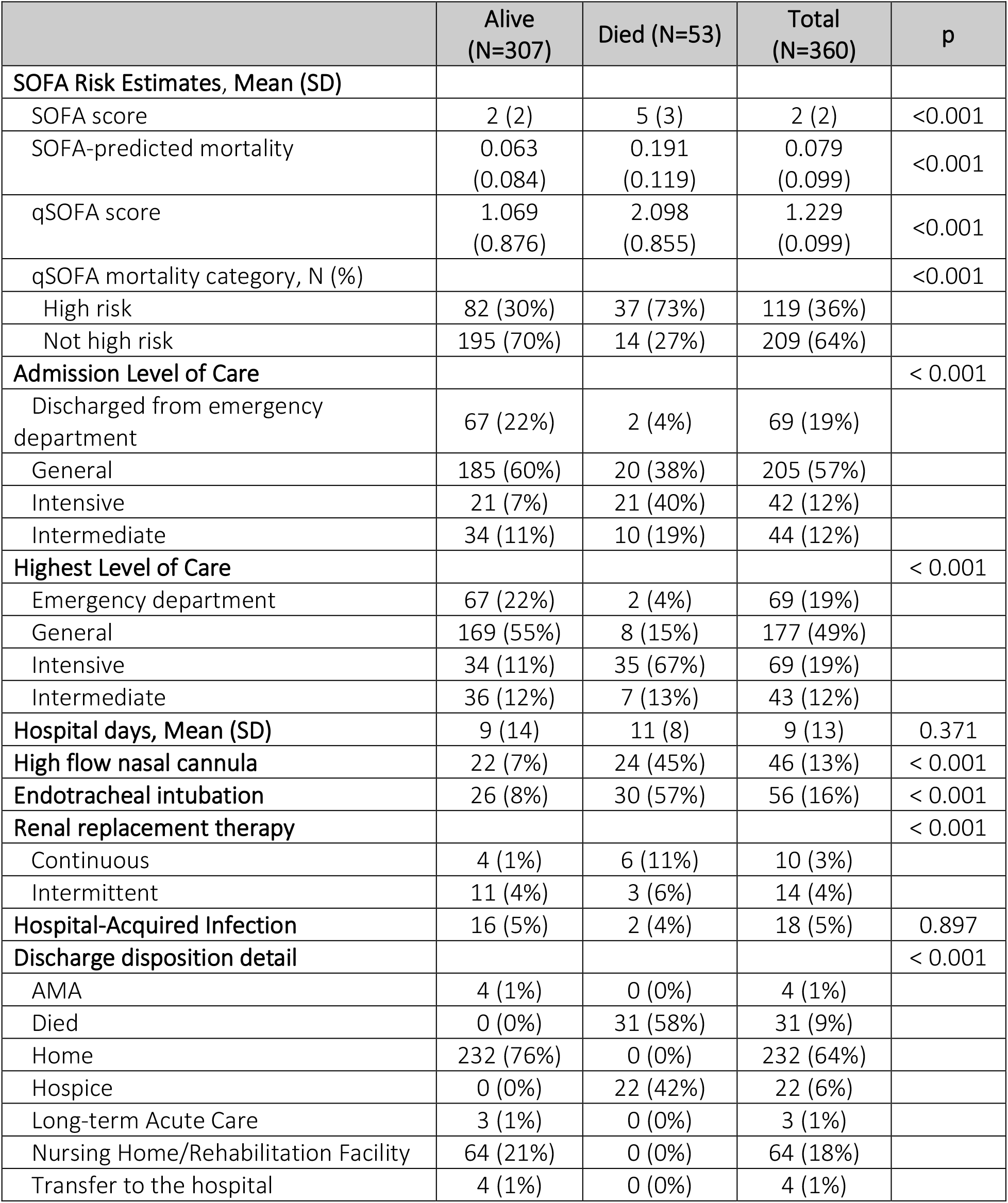
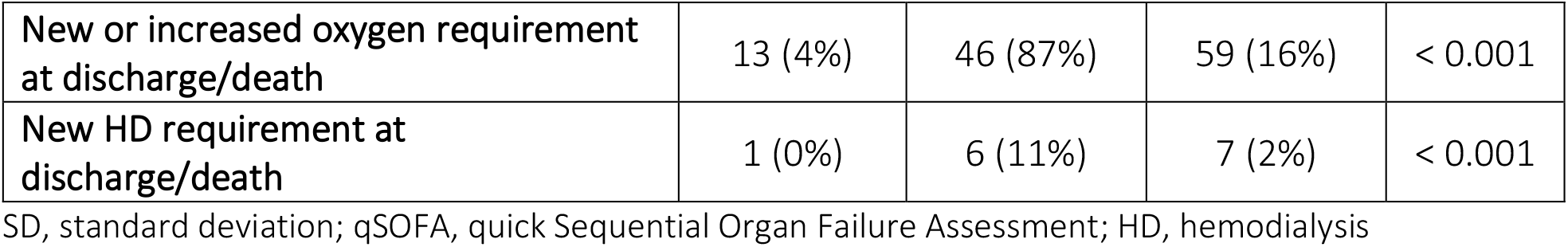
Clinical Characteristics of Hospitalized Patients with COVID-19 by Mortality Status

**Table 3.** Demographic and Medical Characteristics of Hospitalized Patients with COVID-19 by Age Category

|  | <60 (N=190) | ≥60 (N=170) | Total (N=360) | p |
| --- | --- | --- | --- | --- |
| Hypertension | 89 (47%) | 147 (86%) | 236 (66%) | < 0.001 |
| Diabetes mellitus |  |  |  | 0.028 |
| IDDM | 22 (12%) | 25 (15%) | 47 (13%) |  |
| NIDDM | 26 (14%) | 38 (22%) | 64 (18%) |  |
| Congestive heart failure | 18 (9%) | 37 (22%) | 55 (15%) | 0.001 |
| Coronary artery disease | 10 (5%) | 22 (13%) | 32 (9%) | 0.010 |
| Stroke | 19 (10%) | 68 (40%) | 87 (24%) | < 0.001 |
| Dementia | 6 (3%) | 70 (41%) | 76 (21%) | < 0.001 |
| Chronic kidney disease |  |  |  | < 0.001 |
| Yes | 10 (5%) | 30 (18%) | 40 (11%) |  |
| Yes, on dialysis | 9 (5%) | 17 (10%) | 26 (7%) |  |
| Cirrhosis | 2 (1%) | 2 (1%) | 4 (1%) | 0.915 |
| COPD | 6 (3%) | 16 (9%) | 22 (6%) | 0.013 |
| Asthma | 19 (10%) | 9 (5%) | 28 (8%) | 0.096 |
| Structural lung disease | 2 (1%) | 3 (2%) | 5 (1%) | 0.564 |
| OSA/OHS | 9 (5%) | 10 (6%) | 19 (5%) | 0.571 |
| Malignancy |  |  |  | 0.023 |
| Current | 3 (2%) | 8 (5%) | 11 (3%) |  |
| Prior | 2 (1%) | 8 (5%) | 10 (3%) |  |
| HIV |  |  |  | 0.467 |
| Yes | 8 (4%) | 5 (3%) | 13 (4%) |  |
| Radiology |  |  |  |  |
| CXR performed | 174 (92%) | 166 (98%) | 340 (94%) | 0.012 |
| Atelectasis | 45 (26%) | 58 (35%) | 103 (30%) | 0.069 |
| Bilateral Opacities, not effusions | 73 (42%) | 80 (48%) | 153 (45%) | 0.248 |
| No Cavity/abscess | 174 (100%) | 166 (100%) | 340 (100%) | 0.664 |
| Consolidation | 10 (6%) | 14 (8%) | 24 (7%) | 0.334 |
| Interstitial | 15 (9%) | 23 (14%) | 38 (11%) | 0.130 |
| Mass/mass-like | 1 (1%) | 0 (0%) | 1 (0%) | 0.328 |
| Multifocal | 19 (11%) | 35 (21%) | 54 (16%) | 0.010 |
| Pleural effusion | 10 (6%) | 17 (10%) | 27 (8%) | 0.121 |
| Reticulonodular | 4 (2%) | 2 (1%) | 6 (2%) | 0.440 |

\*BMI, body mass index; IDDM, insulin dependent diabetes mellitus; NIDDM, non-insulin dependent diabetes mellitus; COPD, chronic obstructive pulmonary disease; OSA/OHS, obstructive sleep apnea/obesity hypoventilation syndrome; HIV, human immunodeficiency virus

**Table 4.** Clinical Characteristics of Hospitalized Patients with COVID-19 by Age Category

|  | <60 Years-Old (N=190) | ≥ 60 Years-Old (N=170) | Total (N=360) | p |
| --- | --- | --- | --- | --- |
| **Admission Level of Care** |  |  |  | < 0.001 |
| Discharged from emergency department | 61 (32%) | 8 (5%) | 69 (19%) |  |
| General | 102 (54%) | 103 (61%) | 205 (57%) |  |
| Intensive | 10 (5%) | 32 (19%) | 42 (12%) |  |
| Intermediate | 17 (9%) | 27 (16%) | 44 (12%) |  |
| **Highest Level of Care** |  |  |  | < 0.001 |
| Emergency department | 61 (32%) | 8 (5%) | 69 (19%) |  |
| General | 91 (48%) | 86 (51%) | 177 (49%) |  |
| Intensive | 21 (11%) | 48 (29%) | 69 (19%) |  |
| Intermediate | 17 (9%) | 26 (15%) | 43 (12%) |  |
| **Hospital days, Mean (SD)** | 6 (10) | 13 (15) | 9 (13) | < 0.001 |
| **ICU days, Mean (SD)** | 186 (803) | 11 (13) | 64 (443) | 0.132 |
| **SOFA Risk Estimates, Mean (SD)** |  |  |  |  |
| SOFA score | 1 (2) | 3 (2) | 2 (2) | < 0.001 |
| SOFA-predicted mortality | 0.042 (0.076) | 0.113 (0.105) | 0.079 (0.099) | < 0.001 |
| qSOFA score | 1 (1) | 2 (1) | 1 (1) | < 0.001 |
| qSOFA mortality category, N (%) |  |  |  | < 0.001 |
| High risk | 28 (17.8%) | 91 (53.2%) | 119 (36.3%) |  |
| Not high risk | 129 (82.2%) | 80 (46.8%) | 209 (63.7%) |  |
| **High flow nasal cannula** | 13 (7%) | 33 (20%) | 46 (13%) | < 0.001 |
| **Endotracheal intubation** | 19 (10%) | 37 (22%) | 56 (16%) | 0.002 |
| **Renal replacement therapy** |  |  |  | 0.395 |
| Continuous | 6 (3%) | 4 (2%) | 10 (3%) |  |
| Intermittent | 5 (3%) | 9 (5%) | 14 (4%) |  |
| **Hospital-Acquired Infection** |  |  |  | 0.235 |
| Yes | 8 (4%) | 10 (6%) | 18 (5%) |  |
| Unknown | 9 (5%) | 3 (2%) | 12 (3%) |  |
| **Discharge disposition detail** |  |  |  | < 0.001 |
| AMA | 4 (2%) | 0 (0%) | 4 (1%) |  |
| Died | 7 (4%) | 24 (14%) | 31 (9%) |  |
| Home | 168 (88%) | 64 (38%) | 232 (64%) |  |
| Hospice | 0 (0%) | 22 (13%) | 22 (6%) |  |
| Long-term Acute Care | 1 (1%) | 2 (1%) | 3 (1%) |  |
| Nursing Home/Rehabilitation Facility | 8 (4%) | 56 (33%) | 64 (18%) |  |
| Transfer to the hospital | 2 (1%) | 2 (1%) | 4 (1%) |  |
| **New or increased oxygen requirement at discharge/death** | 12 (6%) | 47 (28%) | 59 (16%) | < 0.001 |
| **New HD requirement at discharge/death** | 5 (3%) | 2 (1%) | 7 (2%) | 0.315 |

SD, standard deviation; qSOFA, quick Sequential Organ Failure Assessment; HD, hemodialysis; AMA,
against medical advice

## DISCUSSION

In this study of clinical outcomes among 360 patients with COVID-19 at a large Southern safety-net hospital during the first wave of the COVID-19 pandemic, we found a high overall death rate of 15% with the majority of deaths (87%) occurring in persons ≥60 years. Furthermore, most deaths occurred among residents of LCFs, many of whom were afflicted with multiple comorbidities. Our findings highlight the vulnerability of elderly populations living in LCFs to communicable disease and stress the need to implement better preventive and control measures to deal with such crises now and in the future.

Our findings also highlight the disproportionate burden of COVID-19 and related death among certain racial groups. We found the majority of patients who were hospitalized (80%) with and died (87%) from COVID-19 were Black. Compared to all patients tested for COVID-19, we found that persons ≥60 years (44 vs. 34.6%) or Black or African American (80 vs. 70.9%) were more likely to be infected with COVID-19. Additionally, a prior report on patients hospitalized with COVID-19 in Georgia found a similar a prevalence of Black patients (83%) which was much higher than the proportion of Black persons among all hospitalized patients (47%) in the same period. ^15^ A large cohort of patients hospitalized with COVID-19 in Louisiana echoed findings in Georgia, with 77% of all hospitalized patients being Black and 71% of deaths occurring among those who were Black, despite the fact that Blacks comprised only 31% of the health population. ^16^ Social vulnerabilities and racial inequalities are likely major factors leading to racial disparities in COVID-19 illness. ^17^ The racial health disparities exposed by the COVID-19 pandemic have created needed opportunity and momentum for the healthcare system to address health inequities and improve the health and well-being of all persons in the US. ^18^

Our high deaths rates among older patients most of whom resided in LCFs was alarming and raises several important points. First, older patients are at much higher risk for COVID-19 related mortality. This has been shown for the US where 81% of all COVID-19 related deaths are among persons 65 and older and in other countries with available data that have similarly found increased death rates with age. ^19, 20^ The reasons underpinning this association are likely multifactorial and include high prevalence of multiple comorbidities and immune dysfunction. In our cohort, high rates of multiple comorbidities including hypertension, diabetes, congestive heart failure, prior stroke and dementia were present in patients’ ≥60 years and were associated with death in bivariate analysis. These comorbidities have been found in other studies to be common among patients with COVID-19 and their association with progression to severe disease has led to their inclusion as criteria for monoclonal antibody therapy and vaccination. ^21^ Additional age related factors related to the increased risk of death in older patients are immunosenescence and inflammaging.^22, 23^ Immunosenescence refers to a gradual deterioration of the immune system with age which can lead to a decreased protective response to infection and vaccination.^24^ Inflammaging is a more recent concept and refers to a chronic proinflammatory state that is related to immunosenescence.^25^ As severe COVID-19 illness is characterized by high levels of proinflammatory cytokines, inflammaging has been proposed as one explanation for the increased risk of severe disease and death among older persons.^26^ Our results demonstrate the presence of more severe disease at presentation with increasing age, as we found persons ≥ 60 years had higher SOFA scores, multifocal disease on chest x-ray, and were more likely to be admitted to the ICU. A recent study also highlights the potential impact of a declining immune system with age. Among a population observational cohort, investigators evaluated rates of reinfection with SARS-CoV-2 and found that protection against repeat infection was only 47% among persons ≥ 65 years versus >80% in the overall cohort. ^27^ This finding highlights the urgent need to study the durability of immunity following vaccination and to evaluate the utility and need for a booster vaccine among older persons.

Our extremely high rate of hospitalizations and death from COVID-19 among residents of LCFs highlights the vulnerability of people residing in such settings to communicable diseases and urges us to do more to prevent this disproportionate burden of disease in LCFs. According to the COVID-Tracking Project, persons living in LCFs represent 1% of the US population, 5% of all COVID-19 cases, and account for an alarming 34% of US COVID-19 deaths with rates varying from 15-69% depending on the state.^28^ A combination of high risk patients and high risk setting are likely responsible for such a high rate of death among LCF residents with COVID-19. As discussed in the above paragraph, increasing age puts LCF residents at higher risk of severe disease. Additionally, older persons in LCFs versus those living in the community are more likely to have multiple comorbidities including physical and cognitive impairments that place them at higher risk of severe disease and death due to COVID-19.^29^ The disproportionate burden of COVID-19 at LCFs has led to increased attention and scrutiny of their care and availability of resources including staffing and personal protective equipment. Pertinent to our findings is a report highlighting the lack of adequate staffing and resource at a local LCF leading to decreased quality of care and high COVID-19 transmission among residents.^30^ The COVID-19 pandemic has made it clear that we need to improve resources for LCFs to enable them to provide improved care for their residents. Evidence to support the need for more resources includes data finding adequate nurse staffing is one of the strongest factors correlated with improved COVID-19 control and strengthening infection control measures and universal testing can decrease COVID-19 related mortality in LCFs. ^31,32^ The high rate of disease and death due to COVID-19 among residents clearly justifies the prioritization of LCF residents and staff for vaccination; however, despite high rates of vaccination reported among residents early rates of vaccination of staff was only 38%.^33^ More needs to be done to increase vaccine update among healthcare staff to prevent disease among LCFs and also to prevent transmission of COVID-19 from LCFs to the community.^34^ Additionally, the pandemic made us more of aware of our responsibility as a society to take care of our elders, and we as healthcare providers recognize that we have our moral imperative to advocate for the health residents of LCF as they cannot often do so themselves and that we need to take more purposeful action. Beyond being the right and ethical course of action, by advocating for and helping implement improved health systems in LCFs, we will be better prepared as a society us to prevent spread of the current COVID-19 pandemic, SARs-CoV-2 variants that arise,^35^ and future respiratory virus pandemics.

### Limitations

This study had important limitations. The demographics of the catchment area of this hospital may limit generalizability of our findings. However, these limitations may be balanced by enrichment for potentially under-represented groups, which may be informative for future COVID-19 health disparities research. Given our study period is limited to the first wave of the pandemic, our high death rate may have been due in part to diagnostic delays and lack of available treatments for severe COVID-19. However, current data indicates death rates remain high among older adults and that this population still contributes to the majority of COVID-19 associated mortality. ^36^

## Conclusion

Our results confirm other reports that the burden of COVID-19 associated death is occurring among older adults living in LCFs, and adds evidence to the growing call for better healthcare infrastructure in such settings. With the expected substantial growth of the ≥ 60 population over the next few decades^37^ it is imperative that we put in place improved preventive and treatment measures to protect our elderly population.

## Data Availability

All data produced in the present study are available upon reasonable request to the authors.

## Notes

**Conflict of Interest Statement:** No authors declare a conflict of interest.

### Competing Interest Statement

The authors have declared no competing interest.

### Funding Statement

This study did not receive any funding.

### Author Declarations

The study was approved by the Emory University Institutional Review Board and the Grady Memorial Hospital Research Oversight Committee.

## REFERENCES

1. Apuzzo M, Kirkpatrick DD, April 1, 2020. Covid-19 Changed How the World Does Science, Together. New York Times

2. Beigel JH, Tomashek KM, Dodd LE, Mehta AK, Zingman BS, Kalil AC, Hohmann E, Chu HY, Luetkemeyer A, Kline S, Lopez de Castilla D, Finberg RW, Dierberg K, Tapson V, Hsieh L, Patterson TF, Paredes R, Sweeney DA, Short WR, Touloumi G, Lye DC, Ohmagari N, Oh MD, Ruiz-Palacios GM, Benfield T, Fatkenheuer G, Kortepeter MG, Atmar RL, Creech CB, Lundgren J, Babiker AG, Pett S, Neaton JD, Burgess TH, Bonnett T, Green M, Makowski M, Osinusi A, Nayak S, Lane HC, Members A-SG, 2020. Remdesivir for the Treatment of Covid-19 - Final Report. N Engl J Med 383: 1813–1826.

3. Jackson LA, Anderson EJ, Rouphael NG, Roberts PC, Makhene M, Coler RN, McCullough MP, Chappell JD, Denison MR, Stevens LJ, Pruijssers AJ, McDermott A, Flach B, Doria-Rose NA, Corbett KS, Morabito KM, O’Dell S, Schmidt SD, Swanson PA, 2nd, Padilla M, Mascola JR, Neuzil KM, Bennett H, Sun W, Peters E, Makowski M, Albert J, Cross K, Buchanan W, Pikaart-Tautges R, Ledgerwood JE, Graham BS, Beigel JH, m RNASG, 2020. An mRNA Vaccine against SARS-CoV-2 - Preliminary Report. N Engl J Med 383: 1920–1931.

4. Team IC-F, 2020. Modeling COVID-19 scenarios for the United States. Nat Med.

5. Rahmandad H, Lim TT, Sterman J, 2020. Behavioral dynamics of COVID-19: estimating under-reporting, multiple waves, and adherence fatigue across 91 nations. medRxiv (Preprint). November 26, 2020. [cited 2020 December 19]. Available from: https://doi.org/10.1101/2020.06.24.20139451.

6. Boehmer TK, DeVies J, Caruso E, van Santen KL, Tang S, Black CL, Hartnett KP, Kite-Powell A, Dietz S, Lozier M, Gundlapalli AV, 2020. Changing Age Distribution of the COVID-19 Pandemic - United States, May-August 2020. MMWR Morb Mortal Wkly Rep 69: 1404–1409.

7. Centers for Disease Control and Prevention. COVID Data Tracker.. Available at: https://covid.cdc.gov/covid-data-tracker/#datatracker-home. Accessed February 12, 2021.

8. 2020. COVID-19 Dashboard by the Center for Systems Science adn Engineering (CSSE) at John Hopkings University

9. McMichael TM, Currie DW, Clark S, Pogosjans S, Kay M, Schwartz NG, Lewis J, Baer A, Kawakami V, Lukoff MD, Ferro J, Brostrom-Smith C, Rea TD, Sayre MR, Riedo FX, Russell D, Hiatt B, Montgomery P, Rao AK, Chow EJ, Tobolowsky F, Hughes MJ, Bardossy AC, Oakley LP, Jacobs JR, Stone ND, Reddy SC, Jernigan JA, Honein MA, Clark TA, Duchin JS, Public H-S, King County E, Team CC-I, 2020. Epidemiology of Covid-19 in a Long-Term Care Facility in King County, Washington. N Engl J Med 382: 2005–2011.

10. Chidambaram P, Garfield R, 2020. COVID-19 Has Claimed the Lives of 100,000 Long-Term Care Residents and Staff Available at: https://www.kff.org/policy-watch/covid-19-has-claimed-the-lives-of-100000-long-term-care-residents-and-staff/?utm_campaign=KFF-2020-Coronavirus&utm_medium=email&_hsmi=100922300&_hsenc=p2ANqtz-_W17kM7RAENqsr6EbfwZb_f4cHbZ5qES41WGYyjYmzhDITTrYmLlYbnnxUP6YRdCrkFrPN0l_0Nm8hzj5oLmBx5eZJ8Q&utm_content=100922300&utm_source=hs_email. Accessed January 4, 2021

11. Woolf SH, Chapman DA, Lee JH, 2020. COVID-19 as the Leading Cause of Death in the United States. JAMA.

12. Szilagyi PG, Thomas K, Shah MD, Vizueta N, Cui Y, Vangala S, Kapteyn A, 2020. National Trends in the US Public’s Likelihood of Getting a COVID-19 Vaccine-April 1 to December 8, 2020. JAMA.

13. Harris PA, Taylor R, Minor BL, Elliott V, Fernandez M, O’Neal L, McLeod L, Delacqua G, Delacqua F, Kirby J, Duda SN, Consortium RE, 2019. The REDCap consortium: Building an international community of software platform partners. J Biomed Inform 95: 103208.

14. Vincent JL, Moreno R, Takala J, Willatts S, De Mendonca A, Bruining H, Reinhart CK, Suter PM, Thijs LG, 1996. The SOFA (Sepsis-related Organ Failure Assessment) score to describe organ dysfunction/failure. On behalf of the Working Group on Sepsis-Related Problems of the European Society of Intensive Care Medicine. Intensive Care Med 22: 707–10.

15. Gold JAW, Wong KK, Szablewski CM, Patel PR, Rossow J, da Silva J, Natarajan P, Morris SB, Fanfair RN, Rogers-Brown J, Bruce BB, Browning SD, Hernandez-Romieu AC, Furukawa NW, Kang M, Evans ME, Oosmanally N, Tobin-D’Angelo M, Drenzek C, Murphy DJ, Hollberg J, Blum JM, Jansen R, Wright DW, Sewell WM, 3rd, Owens JD, Lefkove B, Brown FW, Burton DC, Uyeki TM, Bialek SR, Jackson BR, 2020. Characteristics and Clinical Outcomes of Adult Patients Hospitalized with COVID-19 - Georgia, March 2020. MMWR Morb Mortal Wkly Rep 69: 545–550.

16. Price-Haywood EG, Burton J, Fort D, Seoane L, 2020. Hospitalization and Mortality among Black Patients and White Patients with Covid-19. N Engl J Med 382: 2534–2543.

17. Kim SJ, Bostwick W, 2020. Social Vulnerability and Racial Inequality in COVID-19 Deaths in Chicago. Health Educ Behav 47: 509–513.

18. Lopez L, 3rd, Hart LH, 3rd, Katz MH, 2021. Racial and Ethnic Health Disparities Related to COVID-19. JAMA 325: 719–720.

19. COVID-19 Mortality Mortality Overiew. https://www.cdc.gov/nchs/covid19/mortality-overview.htm. Accessed March 19, 2021.. Accessed.

20. Kang SJ, Jung SI, 2020. Age-Related Morbidity and Mortality among Patients with COVID-19. Infect Chemother 52: 154–164.

21. Gasmi A, Peana M, Pivina L, Srinath S, Gasmi Benahmed A, Semenova Y, Menzel A, Dadar M, Bjorklund G, 2021. Interrelations between COVID-19 and other disorders. Clin Immunol 224: 108651.

22. Nikolich-Zugich J, 2018. The twilight of immunity: emerging concepts in aging of the immune system. Nat Immunol 19: 10–19.

23. Nikolich-Zugich J, Knox KS, Rios CT, Natt B, Bhattacharya D, Fain MJ, 2020. SARS-CoV-2 and COVID-19 in older adults: what we may expect regarding pathogenesis, immune responses, and outcomes. Geroscience 42: 505–514.

24. Aw D, Silva AB, Palmer DB, 2007. Immunosenescence: emerging challenges for an ageing population. Immunology 120: 435–46.

25. Fulop T, Larbi A, Witkowski JM, 2019. Human Inflammaging. Gerontology 65: 495–504.

26. Robinson LA, Pierce CM, 2020. Is ‘inflammaging’ fuelling severe COVID-19 disease? J R Soc Med 113: 346–349.

27. Hansen CH, Michlmayr D, Gubbels SM, Molbak K, Ethelberg S, 2021. Assessment of protection against reinfection with SARS-CoV-2 among 4 million PCR-tested individuals in Denmark in 2020: a population-level observational study. Lancet.

28. The Long-Term Care COVID Tracker. https://www.kff.org/coronavirus-covid-19/issue-brief/state-covid-19-data-and-policy-actions/#longtermcare. Accessed March 24, 2021.. Accessed.

29. Panagiotou OA, Kosar CM, White EM, Bantis LE, Yang X, Santostefano CM, Feifer RA, Blackman C, Rudolph JL, Gravenstein S, Mor V, 2021. Risk Factors Associated With All-Cause 30-Day Mortality in Nursing Home Residents With COVID-19. JAMA Intern Med.

30. Low nursing home staffing puts residens at risk. Atlanta Journal Constitution. https://www.ajc.com/news/investigations/low-nursing-home-staffing-puts-residents-at-risk/DM6ZMICKVVAG7D3PKBO6ZXRMEM/. Accessed MArch 23, 2021.. Accessed.

31. Figueroa JF, Wadhera RK, Papanicolas I, Riley K, Zheng J, Orav EJ, Jha AK, 2020. Association of Nursing Home Ratings on Health Inspections, Quality of Care, and Nurse Staffing With COVID-19 Cases. JAMA 324: 1103–1105.

32. McBee SM, Thomasson ED, Scott MA, Reed CL, Epstein L, Atkins A, Slemp CC, 2020. Notes from the Field: Universal Statewide Laboratory Testing for SARS-CoV-2 in Nursing Homes - West Virginia, April 21-May 8, 2020. MMWR Morb Mortal Wkly Rep 69: 1177–1179.

33. Gharpure R, Guo A, Bishnoi CK, Patel U, Gifford D, Tippins A, Jaffe A, Shulman E, Stone N, Mungai E, Bagchi S, Bell J, Srinivasan A, Patel A, Link-Gelles R, 2021. Early COVID-19 First-Dose Vaccination Coverage Among Residents and Staff Members of Skilled Nursing Facilities Participating in the Pharmacy Partnership for Long-Term Care Program - United States, December 2020-January 2021. MMWR Morb Mortal Wkly Rep 70: 178–182.

34. Chen MK, Chevalier JA, Long EF, 2021. Nursing home staff networks and COVID-19. Proc Natl Acad Sci U S A 118.

35. Krutikov M, Hayward A, Shallcross L, 2021. Spread of a Variant SARS-CoV-2 in Long-Term Care Facilities in England. N Engl J Med.

36. Chidambaram P, Garfield R, 2021. Patterns in COVID-19 Cases and Deaths in Long-Term Facilities in 2020. Available at: https://www.kff.org/coronavirus-covid-19/issue-brief/patterns-in-covid-19-cases-and-deaths-in-long-term-care-facilities-in-2020/. Accessed February 12, 2021.

37. Mark Mather, Linda A. Jacobsen, and Kelvin M. Pollard, “Aging in the United States,” Population Bulletin 70, no. 2 (2015).

